# Levels and predictors of knowledge, attitude and practice of contraception among female TV undergraduates in Nigeria: a cross-sectional study

**DOI:** 10.1101/2024.01.01.24300698

**Authors:** Hadizah Abigail Agbo, Philip Adewale Adeoye, Danjuma Ropzak Yilzung, Jawa Samson Mangut, Paul Friday Ogbada

## Abstract

**Introduction:** Access to contraception is a preventive measure against unplanned pregnancy and sexually transmitted infections; especially in sub-Saharan Africa where unmet need is a huge public health concern. This study assessed the levels and predictors of knowledge, attitude and practice of contraception among female TV studies students in Nigeria.

**Methods:** This is a cross-sectional study was conducted among female students of NTA TV College, Nigeria. Predictors of good knowledge, attitude and practice of contraception were determined by multivariable binary logistic regression. A p-value of < 0.05 was determined to be statistically.

**Result:** There were 217 study participants; with an average age of 22 years. Levels of good knowledge, attitude and practice of contraception were 55.3%, 47.5%, and 50.7% respectively. About 74% has ever had sex; with their main sources of contraceptive information from friends and the internet; and commonly used contraceptives are condom and oral contraceptive pills. The commonest reason for the non-use of contraceptives is fear of side effects or health risks. Being a young adult is a significant predictor (aOR: 2.633 [95% CI: 1.031-6.723); p=0.043) of good knowledge; while being a diploma student (aOR: 2.369 [95%CI: 1.219-4.604]; p=0.011); living off-campus (aOR: 2.134 [95%CI: 1.043-4.368]; p=0.038) and good knowledge (aOR: 3.798 [95%CI: 2.079-6.937]; p=0.000) are significant predictors of good attitude. Being from the state’s indigenous population (aOR: 2.369 [95%CI: 1.222-4.595]; p=0.011) and ever engaging in sex (aOR: 24.482 [95%CI: 7.914-75.736]; p=0.000) are significant predictors of good contraceptive use.

**Conclusion:** Our study has shown relatively low levels of good knowledge, attitude and practice of contraception and their predictors. Therefore, there is an urgent need to improve advocacy and policies to improve knowledge, attitude and practice on contraception and sexual and reproductive health services among young people.

## INTRODUCTION

Globally, the proportion of women with unmet need for modern contraception is highest in sub-Saharan Africa-twice the world’s average.^1^ These unmet need has been reported to lead to unwanted pregnancy, unsafe abortion and the limited ability of women to advance educationally, career-wise and economically. The use of contraceptives should be a right-based issue that is necessary for ensuring informed choices regarding family planning. Correct use can significantly improve women’s reproductive health and well-being.^2^ Contraception can be said to be an important measure of unintended pregnancy, abortion and STI, especially the young people. Recent efforts, in the last decade, have sought to reduce unmet needs among women and girls.^3^

The proportion of youths in Nigeria is reported to be about half of the population; with about 57% never married.^4^ This group constitute the highest proportion of those that join the higher education systems yearly. Though this group is the most sexually active; utilization is higher, but the highest level of unmet need among all population groups.^5–8^ They are more at risk of pre-marital sex -often without protection; early, multiple but short-lived intimate relationships; limited knowledge on sexuality issues which is required for healthy sex life and reduction in the risk of teenage pregnancy; unsafe abortion, STIs among others; and less likely to discuss family planning issues with healthcare providers^6,8^; which poses public health and social problems in many LMICs; with many studies indicating increasing incidence of unsafe abortion, STIs including HIV/AIDS, violence against girls, pregnancy-related morbidity and mortality.^8–10^ Unintended and unwanted pregnancy among university students may jeopardize their academic pursuit and potential future career.

Despite the increased accessibility of contraceptives at health facilities across the country, utilization remains low among young females. General uptake variations across the region with the northcentral one of the highest with unmet needs. Girls faced more challenges accessing contraceptives than women living with intimate partners due to associated stigma associated with their pre-marital sexual activities.^1,6,8,11^ Also, they are also faced with limited access to healthful information about contraception – regional variations; with their perception being guided by the prevailing socio-cultural norms and peer-influence.^1,6,8^ Thus, young people lack the needed self-efficacy to negotiate a healthy sexual encounter.

Several studies have reported the contraceptive knowledge attitude and practice among various groups of young people. Recent reports have shown that exposure to mass media communication regarding family planning increases the likelihood of usage in sub-Saharan Africa.^12^ With Nigeria’s high fertility rate and a corresponding maternal mortality rate^6,13,14^; efforts should be geared at increasing and monitoring access to family planning information and services among young women to ensure a healthy reproductive life and general well-being.

However, little is known about contraceptive knowledge, attitude and practice among tertiary education students in mass communication, journalism and TV studies disciplines; as this group of students may be involved in the conceptualization, development and implementation of ICE and mass media campaign activities regarding contraception among communities and the nation in the future. Also, the state where the study was done has been shown to have the highest level of unmet need in Nigeria’s northcentral region among married females^6^; and therefore a need to assess contraceptive behaviour among the study population. This study assessed the level and predictors of knowledge, attitude and practice of contraception among female TV studies students in Nigeria.

## Methods

The conceptual theory for this study is the Health Belief Model. There are 7 constructs in the model and these constructs were applied to the utilization of contraceptives among the study participants. Perceived susceptibility to unwanted pregnancy may make an individual evaluate the perceived severity or consequences of unintended pregnancy. This may drive the individual to evaluate the perceived benefits of contraceptive use to prevent the perceived threat of contraceptive non-use. Perceived barriers to contraceptive use will be thoroughly evaluated to decide on the feasibility of contraceptive use. However, because human behavioural change takes time, there will be the need to remind individuals to adopt and maintain contraceptive use often (cues to action) via mass media; information and communication technology, family and peers; and the development of self-efficacy for contraceptive use.^15^

### Study area

Plateau State is located in the North Central part of Nigeria. It covers a land area of 26,899 square kilometers; with an estimated population of about 4.9 million.^16^ It has over 40 ethnolinguistic groups and each group has its distinct language. English and Hausa are common spoken languages in Plateau State. It is bounded in the northeast by Bauchi State, in the north-west by Kaduna; in the south-west by Nasarawa and in the south-east by Taraba state. Though situated in a tropic area, it has a near-temperate climate due to its high altitude. It has 17 local governments in which Jos North, Jos South and Jos East make up the Jos metropolis. It has 12 higher institutions awarding various post-secondary school certificates; among which are 5 specialized educational institutions such as NTA TV college.^16^

NTA TV College was created, in 1980, to meet the need to train the TV workforce to meet broadcast challenges in Nigeria. It is located in Rayfield, Jos South LGA, Jos, Plateau State. At the earliest stage, it was concerned with the conduct of continuous professional development and short courses for TV industry stakeholders. It was later upgraded to offer diploma and degree programmes. The college is currently affiliated with the Ahmadu Bello University, Zaria-Nigeria, to offer a Mass Communication degree in TV production and Journalism.^17^

#### Study design

This is a cross-sectional study.

#### Study population

The study population are female Students of NTA Television College Jos, who gave their consent to participate in the study, were included. Female students of NTA Television College Jos who withdraw their consent to participate at any stage of the study; and also, those who are not available for one reason or the other during the research will be excluded from the study.

#### Sample size determination

Calculation of sample size was determined using Cochran’s sample size determination formula^18^; mathematically expressed as:

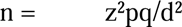

Where, n = Minimum sample size; z = Standard normal deviate at 95% confidence interval = 1.96 p = Proportion of females’ undergraduates’ awareness of contraception= 0.84^19^; q = alternate probability = 1 – 0.84= 0.16; d = Precision = 5% = 0.05. Therefore, n = 206.; adjusting for 10% non-response will make the total sample size to be 227.

#### Sampling technique

The female students were selected using a simple random technique by balloting from the 6 levels during classes in the college.

#### Study instrument/ data collection methods

Data was collected from the female students of NTA Television College Jos by the research team using a semi-structured self-administered questionnaire after informed written consent was obtained. The questionnaire is divided into four sections: social demographic characteristics; Knowledge of contraception; Attitudes to contraceptives; Practice of contraception and Sexual behaviour. The questionnaire was pretested and study piloted among female students of National Film Institute Jos, which are comparable population to our study population to identify errors, test the fidelity of the research process, the feasibility of the study and observe understandability of the research tools by the research participants during the pilot.

#### Data management and analysis

The data obtained were entered and analyzed using Statistical Product and Service Solutions (IBM, Armonk, New York, USA). Qualitative data such as sex and religion were presented using frequencies and percentages, while quantitative data were presented using means and standard deviation-except if not normally distributed. The average scores were used to compute the levels of knowledge, attitude and practice of contraception; with scores less than the average score classified as poor and good level at least the average score. Simple logistic regression was used to determine the factor that is associated with the practice of contraception and to determine the candidate variables for multivariable analysis. Variables that are less than 10% probability were selected and added into the omnibus model to determine the predictors of knowledge, attitude and practice of contraception. A p-value < 0.05 was adjudged significant.

#### Ethical considerations

Ethical clearance was obtained from the JUTH Research and Ethics Committee (JUTH/DCS/IREC/127/XXXI/2619). Written informed consent was obtained before participation; which was voluntary and clients were free to withdraw consent at any point. There was no potential hazard to the participants.

## RESULTS

There was a 96% response rate among study participants.

**Table 1** (below) shows that the majority were young adults with no intimate partner; of the Christian faith; with about two-thirds being outside-of-state students; plateau indigenous; in degree programmes; in TV journalism and more than three-quarters live off-campus. Less than half of the respondents receive at least the minimum wage as a monthly allowance.

**Table 1:** Socio-demographic characteristics of study respondents.

| Variable | Frequency (%) |
| --- | --- |
| **Age** |  |
| ≤ 19 years (teenagers) | 34 (15.7) |
| > 19 years (young adults) | 183 (84.3) |
| *Mean (± SD) 21.9 (± 2.64)* |  |
| **Marital status** |  |
| Not living with a spouse/partner | 198 (91.2) |
| living with a spouse/partner | 19 (8.8.) |
| **Religion** |  |
| Christianity | 186 (85.7) |
| Islam | 31 (14.3) |
| **Tribe/ethnicity** |  |
| Plateau indigenous | 91 (41.9) |
| Plateau non-indigenous | 126 (58.1) |
| **Programme** |  |
| Diploma | 91 (41.9) |
| Degree | 126 (58.1) |
| **Level in school** |  |
| OND1/OND2 (early classes) | 91 (41.9) |
| 100L/200L (middle classes) | 45 (20.7) |
| 300L/400L (older classes) | 81 (37.3) |
| **Monthly income<sup>\$</sup>** | |
| < 18,000 (< \$43.28) | 76 (51.0) |
| ≥ 18,000 (≥ \$43.28) | 73 (49.0) |
| *Median [IQR] ₦17,500 [23,000 – 10000]* |  |
| **Home residence** |  |
| Plateau state | 126 (62.7) |
| Outside Plateau state | 75 (37.3) |
| **School residence** |  |
| Campus | 50 (23.3) |
| Off-campus | 165 (76.7) |
| **Department** |  |
| TV journalism | 149 (69.6) |
| TV production | 65 (30.4) |

<sup>\$</sup>=exchange rate at \$1=₦415 as at February 18, 2022.

**Table 2** (below) shows just above half reported good knowledge, attitude and practice. Almost three-quarters (73.7%) have had sexual intercourse.

**Table 2:** Level of knowledge on, attitude to, practice of contraception and engagement in sexual activity among study respondents.

| Variables | Frequency (percentage) |
| --- | --- |
| **Knowledge level** |  |
| Poor knowledge) | 97 (44.7) |
| Good knowledge) | 120 (55.3) |
| Average Knowledge Score (Median [IQR]) 50.0% [58.0 – 33.0] |  |
| **Attitude level** |  |
| Good attitude | 103 (47.5) |
| Poor attitude | 114 (52.5) |
| Average attitude score Mean (±SD): 71.30% (± 10.63) | |
| **Practice level** |  |
| Poor practice | 107 (49.3) |
| Good practice | 110 (50.7) |
| Average practice score Median [IQR]: 35.0 [52.0 – 28.0] |  |
| **Sexual behaviour** |  |
| Ever had sexual intercourse | 160 (73.7) |
| Never had sexual intercourse | 57 (26.3) |

Figure 1 (below) shows that friends (35.2%) and the internet (34.3%) are the commonest sources of information on contraception.

**Figure 1:**
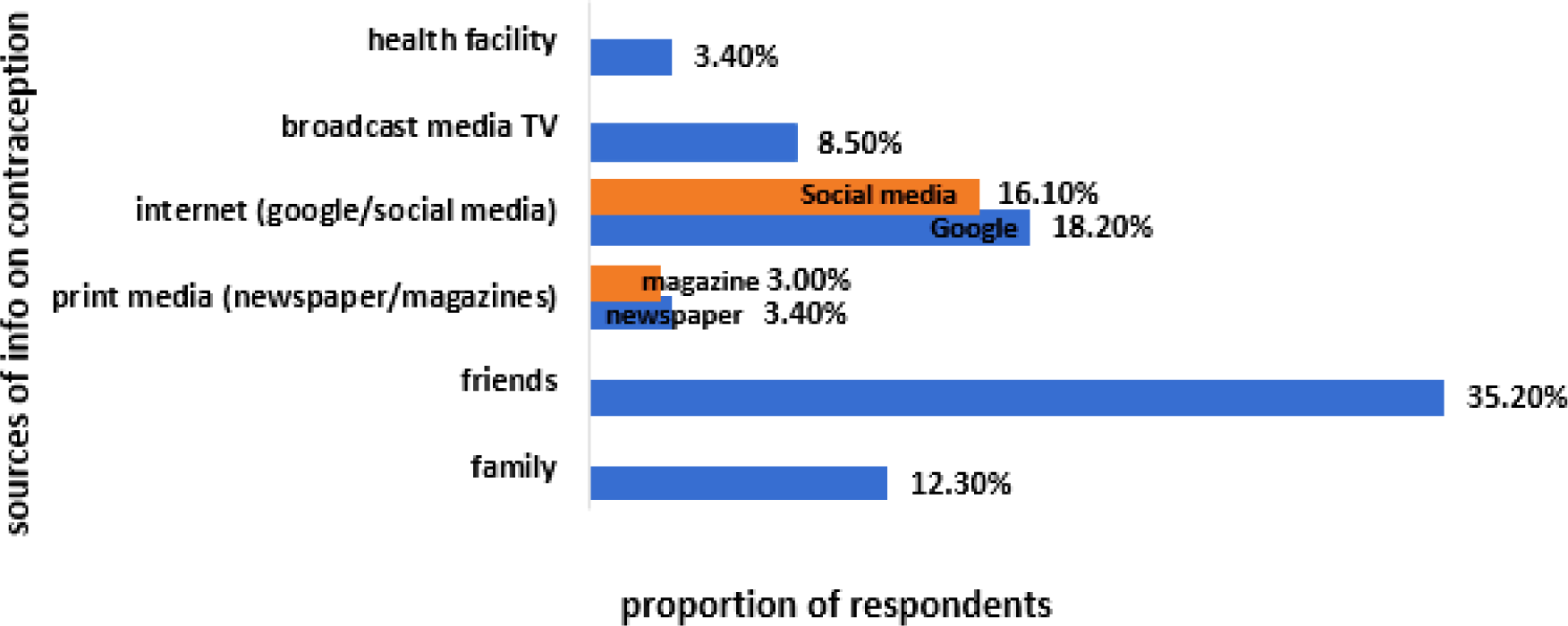
Sources of information on contraception among study respondents.

Figure 2 (below) shows that condoms (43.5%) and oral contraceptive pills (OCPs) (36.5%) are the commonest contraceptives used by students at NTA TV college.

**Figure 2:**
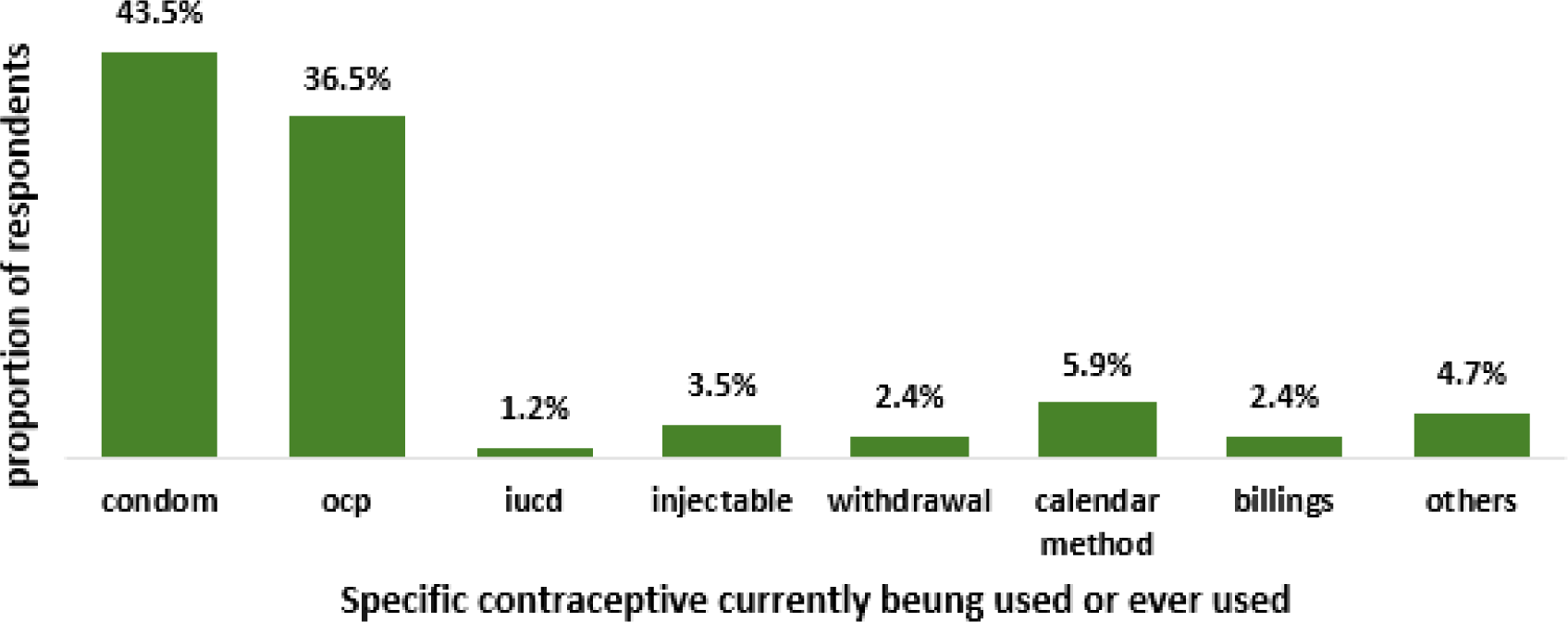
Specific contraceptive currently being used or ever used among study respondents.

Figure 3 (below) shows that the commonest reason why respondents will not use contraceptives is because of side effects.

**Figure 3:**
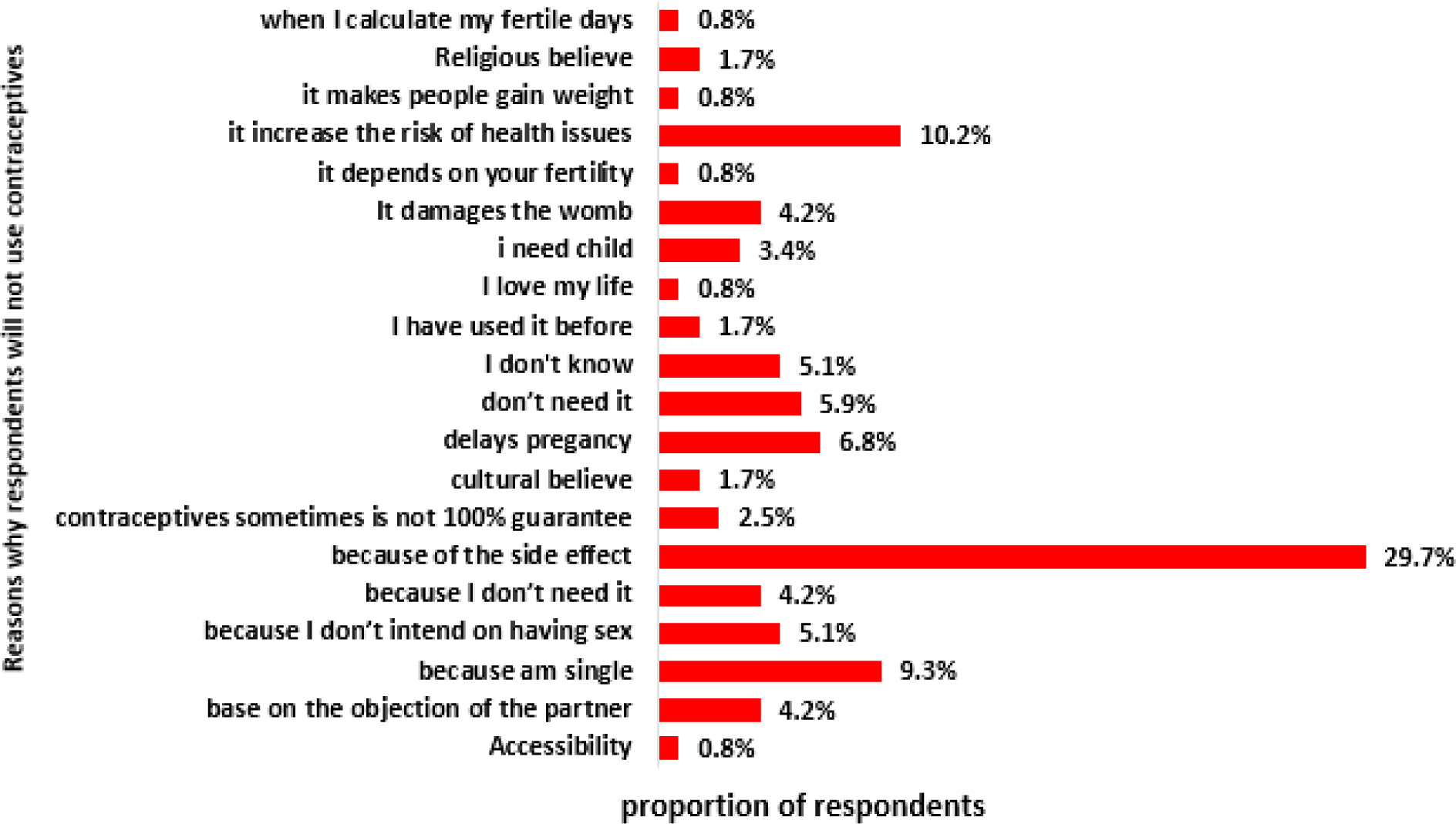
reasons why respondents will not use contraceptives among study respondents.

**Table 3** (below) shows that being a young adult (>19years) is a significant predictor of knowledge on contraception (aOR: 2.633 [95% CI: 1.031-6.723]; p=0.043) compared to teens (≤19 years).

**Table 3:**
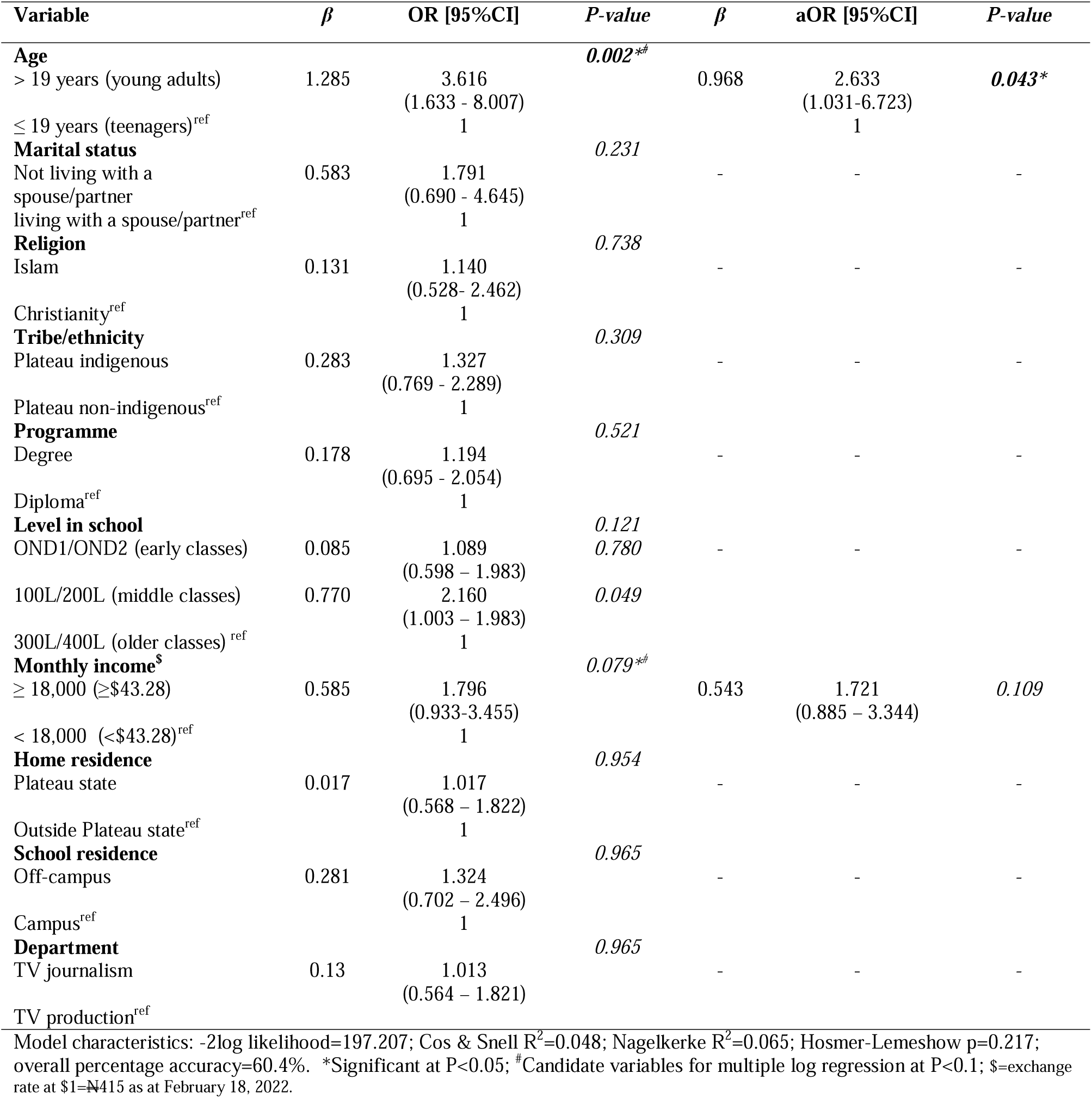
Predictors of contraception knowledge among study respondents.

**Table 4** (below) shows being a diploma student (aOR: 2.369 [95%CI: 1.219-4.604]; p=0.011) is a significant predictor of good attitude to contraception compared to those in degree programmes; having an off-campus accommodation at school is a significant predictor of good attitude (aOR: 2.134 [95%CI: 1.043-4.368]; p=0.038) compared to those on on-campus accommodation; and having good knowledge is a significant predictor of good attitude (aOR: 3.798 [95%CI: 2.079-6.937]; p=0.000) compare to those with poor knowledge.

**Table 4:** Predictors of attitude to contraception among study respondents.

| Variable | $\beta$ | OR [95%CI] | P-value | $\beta$ | aOR [95%CI] | P-value |
| --- | --- | --- | --- | --- | --- | --- |
| **Age** |  |  |  |  |  |  |
| ≤ 19 years (teenagers) | 0.401 | 1.493<br>(0.715 – 3.119) | 0.286 | - | - | - |
| > 19 years (young adults)<sup>ref</sup> |  | 1 |  |  |  |  |
| **Marital status** |  |  |  |  |  |  |
| Not living with a spouse/partner | 0.478 | 1.613<br>(0.610 – 4.269) | 0.335 | - | - | - |
| living with a spouse/partner<sup>ref</sup> |  | 1 |  |  |  |  |
| **Religion** |  |  |  |  |  |  |
| Islam | 0.194 | 1.214<br>(0.567 – 2.598) | 0.618 | - | - | - |
| Christianity<sup>ref</sup> |  | 1 |  |  |  |  |
| **Tribe/ethnicity** |  |  |  |  |  |  |
| Plateau indigenous | 0.137 | 1.147<br>(0.668 – 1.968) | 0.619 | - | - | - |
| Plateau non-indigenous<sup>ref</sup> |  | 1 |  |  |  |  |
| **Programme** |  |  |  |  |  |  |
| Diploma | 0.751 | 2.120<br>(1.225 – 3.669) | **0.007\*<sup>#</sup>** | 0.863 | 2.369<br>(1.219 – 4.604) | **0.011\*** |
| Degree<sup>ref</sup> |  | 1 |  |  | 1 |  |
| **Level in school** |  |  |  |  |  |  |
| 300L/400L (older classes) | -0.707 | 0.493<br>(0.268 – 0.906) | **0.023\*<sup>#</sup>** | - | - | - |
| 100L/200L (middle classes) | -0.832 | 0.435<br>(0.209 – 0.906) | **0.026\*<sup>#</sup>** | -0.263 | 0.769<br>(0.343 – 1.720) | 0.522 |
| OND1/OND2 (early classes)<sup>ref</sup> |  | 1 | **0.026\*<sup>#</sup>** |  | 1 |  |
| **Monthly income<sup>\$</sup>** | | | | | | |
| ≥ 18,000 (≥ \$43.28) | 0.456 | 1.577<br>(0.826 – 3.013) | 0.168 | - | - | - |
| < 18,000(< \$43.28)<sup>ref</sup> | | 1 | | | | |
| **Home residence** |  |  |  |  |  |  |
| Plateau state | 0.177 | 1.193<br>(0.669 – 2.129) | 0.550 | - | - | - |
| Outside Plateau state<sup>ref</sup> |  | 1 |  |  |  |  |
| **School residence** |  |  |  |  |  |  |
| Off-campus | 0.724 | 2.062<br>(1.066 – 3.990) | **0.032\*<sup>#</sup>** | 0.758 | 2.134<br>(1.043 – 4.368) | **0.038\*** |
| Campus<sup>ref</sup> |  | 1 |  |  | 1 |  |
| **Department** |  |  |  |  |  |  |
| TV journalism | 0.060 | 1.062<br>(0.592 – 1.904) | 0.840 | - | - | - |
| TV production<sup>ref</sup> |  | 1 |  |  |  |  |
| **Level of knowledge** |  |  |  |  |  |  |
| Good knowledge (≥ 50%) | 1.244 | 3.469<br>(1.971 – 6.106) | **0.000\*<sup>#</sup>** | 1.334 | 3.798<br>(2.079 – 6.937) | **0.000\*** |
| Poor knowledge (< 50%)<sup>ref</sup> |  | 1 |  |  | 1 |  |

Model characteristics: -2log likelihood=264.244; Cos & Snell R<sup>2</sup>=0.143; Nagelkerke R<sup>2</sup>=0.191; Hosmer-Lemeshow p=0.899; overall percentage accuracy=67.9%. \*Significant at P<0.05; <sup>#</sup>Candidate variables for multiple log regression at P<0.1; \$=exchange rate at \$1=₦415 as at February 18, 2022.

**Table 5** (below) shows that being from the state’s indigenous (majority) population is a significant predictor of good contraceptive practice (aOR: 2.369 [95%CI: 1.222-4.595]; p=0.011) compared to those from the non-indigenous population. Ever engaging in sex (aOR: 24.482 [95%CI: 7.914-75.736]; p=0.000) is a significant predictor of good contraceptive practice compared to those who had never engaged in sex.

**Table 5:** Predictors of practice of contraception among study respondents.

| Variable | $\beta$ | OR [95%CI] | P-value | $\beta$ | aOR [95%CI] | P-value |
| --- | --- | --- | --- | --- | --- | --- |
| **Age** |  |  |  |  |  |  |
| > 19 years (young adults) | 0.748 | 2.114<br>(0.988-4.524) | 0.054<sup>#</sup> | -0.387 | 0.679<br>(0.217-2.126) | 0.506 |
| ≤ 19 years (teenagers)<sup>ref</sup> |  | 1 |  |  | 1 |  |
| **Marital status** |  |  |  |  |  |  |
| living with a spouse/partner | 0.085 | 1.089<br>(0.424-2.795) | 0.859 | - | - | - |
| Not living with a spouse/partner<sup>ref</sup> |  | 1 |  |  |  |  |
| **Religion** |  |  |  |  |  |  |
| Christianity | 0.108 | 1.114<br>(0.520-2.383) | 0.782 | - | - | - |
| Islam<sup>ref</sup> |  | 1 |  |  |  |  |
| **Tribe/ethnicity** |  |  |  |  |  |  |
| Plateau indigenous | 0.997 | 2.711<br>(1.551-4.739) | 0.000*<sup>#</sup> | 0.863 | 2.369<br>(1.222-4.595) | 0.011* |
| Plateau non-indigenous<sup>ref</sup> |  | 1 |  |  | 1 |  |
| **Programme** |  |  |  |  |  |  |
| Degree | 0.161 | 1.175<br>(0.685-2.016) | 0.558 | - | - | - |
| Diploma<sup>ref</sup> |  | 1 |  |  |  |  |
| **Level in school** |  |  |  |  |  |  |
| OND1/OND2 (early classes) | 0.157 | 1.170<br>(0.642-2.134) | 0.608 | - | - | - |
| 100L/200L (middle classes) | 0.916 | 2.500<br>(1.170-5.341) | 0.018*<sup>#</sup> |  |  |  |
| 300L/400L (older classes)<sup>ref</sup> |  | 1 | 0.053<sup>#</sup> |  |  |  |
| **Monthly income<sup>\$</sup>** | | | | | | |
| ≥ 18,000 (≥ \$43.28) | 0.192 | 1.212<br>(0.637-2.308) | 0.558 | - | - | - |
| < 18,000 (< \$43.28)<sup>ref</sup> | | 1 | | | | |
| **Home residence** |  |  |  |  |  |  |
| Outside Plateau state | 0.464 | 1.590<br>(0.890-2.841) | 0.117 | - | - | - |
| Plateau state<sup>ref</sup> |  | 1 |  |  |  |  |
| **School residence** |  |  |  |  |  |  |
| Campus | 0.068 | 1.070<br>(0.568-2.016) | 0.833 | - | - | - |
| Off-campus<sup>ref</sup> |  | 1 |  |  |  |  |
| **Department** |  |  |  |  |  |  |
| TV journalism | 0.337 | 1.401<br>(0.780-2.516) | 0.259 | - | - | - |
| TV production<sup>ref</sup> |  | 1 |  |  |  |  |
| **Level of knowledge** |  |  |  |  |  |  |
| Good knowledge | 0.924 | 2.519<br>(1.454-4.364) | 0.001*<sup>#</sup> | 0.601 | 1.825<br>(0.908-3.668) | 0.091 |
| Poor knowledge<sup>ref</sup> |  | 1 |  |  | 1 |  |
| **Attitude** |  |  |  |  |  |  |
| Good attitude | 0.732 | 2.079<br>(1.209-3.576) | 0.008*<sup>#</sup> | 0.451 | 1.570<br>(0.791-3.116) | 0.197 |
| Poor attitude<sup>ref</sup> |  | 1 |  |  | 1 |  |
| **Sexual behavior** |  |  |  |  |  |  |
| Ever | 3.258 | 26.009<br>(8.941-75.661) | 0.000*<sup>#</sup> | 3.198 | 24.482<br>(7.914-75.736) | 0.000* |
| Never<sup>ref</sup> |  | 1 |  |  | 1 |  |

Model characteristics: -2log likelihood=216.338; Cos & Snell R<sup>2</sup>=0.322; Nagelkerke R<sup>2</sup>=0.43; Hosmer-Lemeshow p=0.992; overall percentage accuracy=75.6%.
\*Significant at P<0.05; <sup>#</sup>Candidate variables for multiple log regression at P<0.1; \$=exchange rate at \$1=₦415 as at February 18, 2022.

## DISCUSSION

Our study shows that about half of all respondents have good knowledge, attitude and practice of contraception among students of TV students with almost three-quarters having ever had sex; with their main sources of contraceptive information being those from friends and the internet; while commonly used contraceptives being condom and OCPs. Non-use of contraceptives is common as a result of fear of side-effect or health risks. Age was observed to be a significant predictor of good knowledge on contraception; while being in a diploma (lower degree); living off-campus and good knowledge are significant predictors of a good attitude to contraception. Ethnicity and sexual behaviour are significant predictors of good contraceptive use.

Our study revealed that about half of respondents have good knowledge. This is similar to a study in Botswana.^20^ However, a lower level of good knowledge is observed among students from Selangor-Malaysia, Spain, Imo state-Nigeria, Adama and emerging regions of Ethiopia;^21–24^ while a higher level of good knowledge is observed among students in Dodoma-Tanzania, Kano and Ilorin-Nigeria, Pretoria-South Africa, Kwadaso-Ghana.^25–29^ Evidence suggests that the introduction of various educational interventions can subsequently increase the level of knowledge.^30^

This study also revealed that almost half of respondents have a good attitude to contraception. This is similar to studies in Selangor-Malaysia and the emerging region of Ethiopia.^21,31^ However, higher levels of good attitude are seen in Kano-Nigeria, Adama and the emerging region of Ethiopia, Pretoria-South Africa, Kwadaso-Ghana and Spain.^22,26,28,29,31^ Thus, non-governmental organizations, healthcare workers and social marketers should introduce strategies to counteract the negative impact of pre-existing beliefs held by young women to increase contraceptive utilization.

It was observed that just half of the study participants have good contraceptive practice. This is lower than that which was reported among students in Kano-Nigeria.^26^ This may be due to the recent 5-state public-private partnership geared towards increasing contraceptive uptake of which Kano is a part. There was also a financial higher commitment to family planning services in these states compared to Plateau state.^32,33^ Therefore, a recent report of the 5-state intervention revealed increased demand generation and uptake, improved state government financing of contraception services.^32^ This government-NGO effort might have rubbed off on young female college students in Kano. Also, recent NDHS reported that women of reproductive age in Kano reported a higher level of exposure to family planning messages and discussion with healthcare workers on family during their visits to health facilities compared to plateau women of reproductive age.^6^

Almost three-quarters have ever had sex. This is similar to sexual behaviour seen among students in Ilorin-Nigeria^27^; lower compared to students in Spain^22^; but higher than those reported among the similar population in Botswana, urban Nigerian cities and Kilimanjaro region of Tanzania.^7,20,34,35^ This may be as a result of an increased liberal worldview among young people; a sense of freedom experienced; and a desire for sexual experimentation in the university environment.

Friends and the internet are the commonest sources of information on contraception. This is similar to studies from Kilimanjaro-Tanzania, Botswana, Ilorin and south-south-Nigeria, Dodoma-Tanzania and Spain among students.^20,22,25,27,34,36^ However, health facilities and healthcare workers are the commonest sources of information on contraception among similar populations in Kwadaso-Ghana and the emerging regions of Ethiopia.^29,31^ Information on contraception from family members often come out of concern that a young person is sexually or about to be sexually active., and therefore the need for safe sex. The family members, sisters and mothers, are highly trusted based on their overall familial relationship. Trust in internet sources is often improved among young women when the source of the information is from reputable sites such as those indicating .org, .edu and .gov.^37^

Condoms and OCPs are the commonest contraceptives among study participants. This is similar to studies from Spain, Dodoma-Tanzania, Botswana, Limpopo-south Africa and south-south Nigeria.^22,25,35,36,38^ This may be a result of their ready availability and accessibility over-the-counter in many jurisdictions. There may be a need to use social marketing approaches to make these contraceptives available to young people to bypass the stigma they experienced while accessing contraceptives from traditional sources of contraceptives.

The commonest reason for contraceptive non-use is the concern for side effects and health risks among the study population. This is similar to studies among similar populations in Botswana, Pretoria-south Africa, Benin republic, Limpopo-South Africa and south-south Nigeria.^11,20,28,36,38^ This might have been driven by personal experiences or information received from significant others such as friends and family members or due to apparent ignorance; even when they’ve never used one; as seen in many developing countries.^1^ Thus, there is a need to individualize contraceptive counselling and choice when encountering young people.

This study shows that being a young adult student is a significant predictor of the acquisition of good knowledge. This is similar to similar national surveys from the US and a study from the emerging regions of Ethiopia.^31,39^ This may be due to less awareness among teenagers and confusing information on contraception online (to which they are more exposed to more than other age groups); their limited capacity to filter, process presented information and make appropriate decisions compared to young adults.^6,7,40^ Therefore early, there is the need for more training in the early training before the onset of sexual relations to prevent the negative consequences of unguarded sexual and reproductive behaviours.

Being a diploma student (lower degree) is a significant predictor of a good attitude to contraception. This is converse to most studies where a higher educational attitude significantly predicts a good attitude to contraception.^1,31^ our result may be due to increased information fatigue following information overload that may occur which might reduce risk perception, and increase nonchalant attitude to contraception. Information received might have been contaminated, over the years with disinformation and misinformation to which higher-level degree students might have been exposed over the years. Thus, information managers and regulators should ensure that information provided is of high value and an opportunity for updates targeted at a specific population without infringing on human rights.

This study shows that being off-campus is a significant predictor of a good attitude. Disaggregation of study data based on school residence shows that off-campus students are older students, married and with higher income, with a higher level of knowledge about contraception compared to on-campus students. Similar demography of off-campus students has been reported among undergraduates in the US.^41^ These demographic characteristics are significant predictors of a good attitude to contraception.^31^

Good knowledge is shown to be a good predictor of a good attitude to contraception. This is similar to studies from emerging regions in Ethiopia and Botswana.^20,31^ Consistently, appropriate and targeted delivery of contraceptive information, education and communication will improve the attitude of specified populations which may further improve contraceptive utilization among young people.

Plateau indigenous (a conglomeration of about 50 ethnic groups and a majority population in the state) is a significant predictor of contraceptive use. This is similar to studies from similar populations from south-south Nigeria, Selangor-Malaysia and the US where being a member of the majority population is a significant predictor of contraceptive use.^21,36,42^ This might be due to disparities in the levels of awareness, knowledge, attitude and access to contraception that has been reported in the majority population. In some instances, minority populations may be wary of the government’s intention to limit minority populations; and also, be sceptic about the safety of government-sanctioned contraceptives.^42,43^ The minority ethnic differences especially in minority populations should spur healthcare providers to provide necessary contraceptive education. Innovative counselling approaches could improve women’s ability to attain an informed decision. Interventions to reach out to tertiary students especially those from minority backgrounds-should be instituted in schools to provide ICE opportunities and students-male and females. Since none of the respondents mentioned any reference to any exposure to sex education in schools,^44^ there may be a need to review the impact of the current sex education policies in schools on the sexual and reproductive health behaviour of young people.

This study shows that being involved in sexual relations is a significant predictor of good contraception practice. This is similar to a study from Kilimanjaro-Tanzania among a similar population.^34^ Individuals who are sexually active see the need to prevent unwanted pregnancy, STIs, pregnancy-related health risks and delay marriage to complete education while in sexual relationships.^9,10,45^ It also helps girls and women achieve empowerment to live a healthy and economically productive life. Studies have shown that sexually inactive young often cite their sexual inactivity as a reason for non-use.^38^

Though the outcome of this study cannot be generalized to all universities, the status of the college being the only TV college in Africa can provide an insight into the KAP of contraception among future TV professionals. There may also be a social desirability bias as respondents might have under-reported sexual behaviours and contraceptive use. There is a need for an appropriate and consistent awareness campaign via acceptable media among university students. Parent-children communication should be encouraged and supported to improve contraceptive KAP; as the family is the first educational institution in the life of children as they prepare to face the world. There is also a need to evaluate and improve on the current comprehensive sexuality education in many sub-Saharan African countries to have more robust training for young people on SRH as early as possible.

## CONCLUSION

Female undergraduate TV College Students in this study relatively have a low level of good knowledge, attitude and practice. There is an urgent need to reform current advocacy efforts, young people-friendly SRH services and policies to improve contraceptive knowledge, attitudes and utilization among young people.

## Data Availability

All data produced in the present study are available upon reasonable request to the authors

## HIGHLIGHTS

- Low level of knowledge and uptake of contraception with unmet need has become a huge public health problem in developing countries.
- There is relatively low levels of good contraceptive knowledge, attitude and practice among study population.
- Age is a significant predictor of good knowledge. Being a diploma student, living off-campus and good knowledge are significant predictors of good contraceptive attitude. Sexual behaviour and being a plateau state indigene are significant predictors of good contraceptive practice.
- There is an urgent need for targeted and appropriate advocacy campaigns, policies and improved young people-friendly SRH services.

## AUTHORS’ CONTRIBUTION

Conceptualization and design: HAO, PAA, DRY, JSM, PFO; Data acquisition: DRY, JSM, PFO; Data analysis and interpretation: PAA; Drafting and critical review: HAO, PAA; Final approval: HAO, PAA, DRY, JSM, PFO.

## ACKNOWLEDGEMENT

We are grateful to the students of NTA TV College of Nigeria for their willingness to participate in the study.

## COMPETING INTEREST

Authors declare no conflict of interest

## FINANACIAL SUPPPORT

Nil

## Notes

### Competing Interest Statement

The authors have declared no competing interest.

### Funding Statement

This study did not receive any funding

### Author Declarations

Ethical clearance was obtained from the JUTH Research and Ethics Committee (JUTH/DCS/IREC/127/XXXI/2619).

